# Coadministration of AYUSH 64 as an adjunct to Standard of Care in mild and moderate COVID-19: A randomised, controlled, multicentric clinical trial

**DOI:** 10.1101/2021.06.12.21258345

**Authors:** Arvind Chopra, Girish Tillu, Kuldeep Chuadhary, Govind Reddy, Alok Srivastava, Muffazal Lakdawala, Dilip Gode, Himanshu Reddy, Sanjay Tamboli, Manjit Saluja, Sanjeev Sarmukkaddam, Manohar Gundeti, Ashwinikumar Raut, BCS Rao, Babita Yadav, Narayanam Srikanth, Bhushan Patwardhan

**Author notes:** **Funding Statement:** This study was funded by Central Council for Research in Ayurvedic Sciences, Ministry of Ayush, Government of India. **Competing interests:** None of the authors have any financial conflict of interest regarding this study. The authors Kuldeep Chuadhary, Alok Srivastava, Govind Reddy, Manohar Gundeti, BCS Rao, Babita Yadav, Narayanam Srikanth work in Central Council for Research in Ayurvedic Sciences (CCRAS), Ministry of AYUSH (MoA), Government of India (GOI), New Delhi. Dr Ashwinikumar Raut was a consultant for the study. Dr Sanjay Tamoli was involved as a CRO. AYUSH-64 is a proprietary formulation of CCRAS. **Contact Address:** Arvind Chopra, MD, Director and Chief Rheumatologist, Centre for Rheumatic Diseases, Pune, India 411001, (M) 9822039297.

## Abstract

**Objectives:** To compare the co-administration of an Ayurvedic drug AYUSH 64 as an adjunct to standard of care (SOC) and SOC for efficacy and safety in the management of COVID-19.

**Design:** Multicentre, parallel efficacy, randomized, controlled, open label, assessor blind, exploratory trial with a convenience sample. Patients followed to complete 12 weeks of study duration.

**Setting:** COVID-19 dedicated non-intensive care wards at 1 government hospital, 1 medical college teaching hospital and 1 medical university teaching hospital

**Participants:** 140 consenting, eligible, hospitalized adult patients suffering from mild and moderate symptomatic COVID-19 and confirmed by a diagnostic (SARS-CoV-2) RT-PCR assay on nasal and throat swab were randomized to SOC or SOC plus AYUSH 64. To be withdrawn if disease becomes severe.

**Interventions:** Two tablets of AYUSH 64, 500 mg each, twice daily after meals, and continued till study completion. SOC (symptomatic and supportive) as per national guidelines of India for mild and moderate disease.

**Main outcome measures:** Time period to clinical recovery (CR) from randomization baseline and proportion with CR within 28 days time frame; CR defined in the protocol

**Results:** 140 patients randomized (70 in each arm); 138 patients with CR qualified for analysis. Both groups were matched at baseline. The mean time to CR from randomization was significantly superior in AYUSH 64 group (95% CI -3.03 to 0.59 days); a higher proportion (69.7%) in the first week (p=0.046, Chi-square). No significant differences observed for COVID-19 related blood assays (such as D-Dimer). AYUSH 64 arm showed significant (p<0.05) superior persistent improvement in general health, quality of life, fatigue, anxiety, stress, sleep and other psychosocial metrics. 1 patient on SOC required critical care. 48 adverse events (AE) reported in each group. Barring three SAE (in SOC), AE were mild and none were drug related. 22 participants (8 on AYUSH) were withdrawn. No deaths were reported.

**Conclusions:** AYUSH 64 hastened recovery, reduced hospitalization and improved overall health in mild and moderate COVID-19 when co-administered with SOC under medical supervision. It was safe and well tolerated. Further studies are warranted.

**Trial registration:** The Clinical Trials Registry India Number CTRI/2020/06/025557

**Funding:** CCRAS, Ministry of AYUSH, Government of India

## INTRODUCTION

The world continues to reel under the tragic burden of the COVID-19 pandemic. The medical system has been precariously overstretched and scarred. Several drug trials were completed and many more are underway to unravel the evidence-based medicine (EBM) for more effective and safe management (1). However, the treatment of mild-moderate COVID-19 remains empirical, symptomatic and supportive (2, 3). Several drugs were repurposed and extensively used for chemoprophylaxis and treatment of COVID-19 (7). However, drugs such as hydroxychloroquine fell into disrepute because of lack of clinical evidence (4). Despite limited evidence but based on good clinical experience, some drugs such as tocilizumab continue to be used (5). Steroids became a pivotal treatment following the result of a single large controlled drug trial (6).

The search to repurpose drugs (COVID-19) also rekindled vigorous research in the traditional, complementary and alternative systems of medicine (TCAM) (8,9). TCAM and herbal medicines in particular can be considered based on available data and clinical experience (10,11). It is now known that the exuberant and dysregulated immune response in COVID-19 leads to life threatening complications. Several medicinal plants from Ayurveda and traditional Chinese medicines which are known to modulate immune response were considered as potential therapeutic candidates (8, 12, 13, 14). Ayurvedic and other traditional herbal medicines are being popularly used in India to prevent and treat COVID-19 since the beginning of the pandemic (15, 16, 17,18). Safety and tolerability of Ayurvedic herbal medicines could be an added advantage for use in the community (19).

India has legal system to regulate and promote plural systems of medicine including Ayurveda, Yoga, Naturopathy, Unani, Siddha, Sowa Rigpa and Homoeopathy, and together are known as AYUSH systems. The Ministry of AYUSH (MoA) has established an Interdisciplinary AYUSH Research and Development Task Force on COVID-19 to promote scientific research (20, 21). Based on Ayurvedic logic and clinical experience, AYUSH-64 was considered for repurpose therapeutic use in COVID-19. It is a standard proprietary poly-herbal formulation of Central Council for Research in Ayurvedic Sciences (CCRAS) which was first developed in 1980 for the treatment of Malaria (22). The current study aims to evaluate AYUSH 64 as an adjunct to standard care in the treatment of COVID-19.

## METHODS

This was a prospective, randomized, open label (assessor blind), two arm multicentre study with an exploratory research design and was planned and carried out during the COVID-19 pandemic (May-Nov 2020). The duration of study was 12 weeks. The study was carried out in the COVID-19 dedicated non-intensive wards in the medical and teaching hospitals at King George Medical University, Lucknow, Central Ayurveda Research Institute for Cancer, Mumbai and Datta Meghe Institute of Medical Sciences, Nagpur. The study was carried out in accordance with the principles of Good Clinical Practice (GCP), Declaration of Helsinki (Brazil update 2013), ICMR (Indian Council of Medical Research) and MoA/CCRAS Guidelines (2018) (23, 24). The protocol and the study report also complied with CONSORT guidelines (25). The protocol was approved by the Institutional Ethics Committee at each study site and registered in the Clinical Trials Registry of India (CTRI) (registration number CTRI/2020/06/025557) prior to patient enrolment (26). The study was monitored by an independent data safety monitoring board and a monitoring committee appointed by the sponsor. The overall scheme of the study, study procedures and predetermined time points of evaluation are shown in Fig 1.

**Figure 1:**
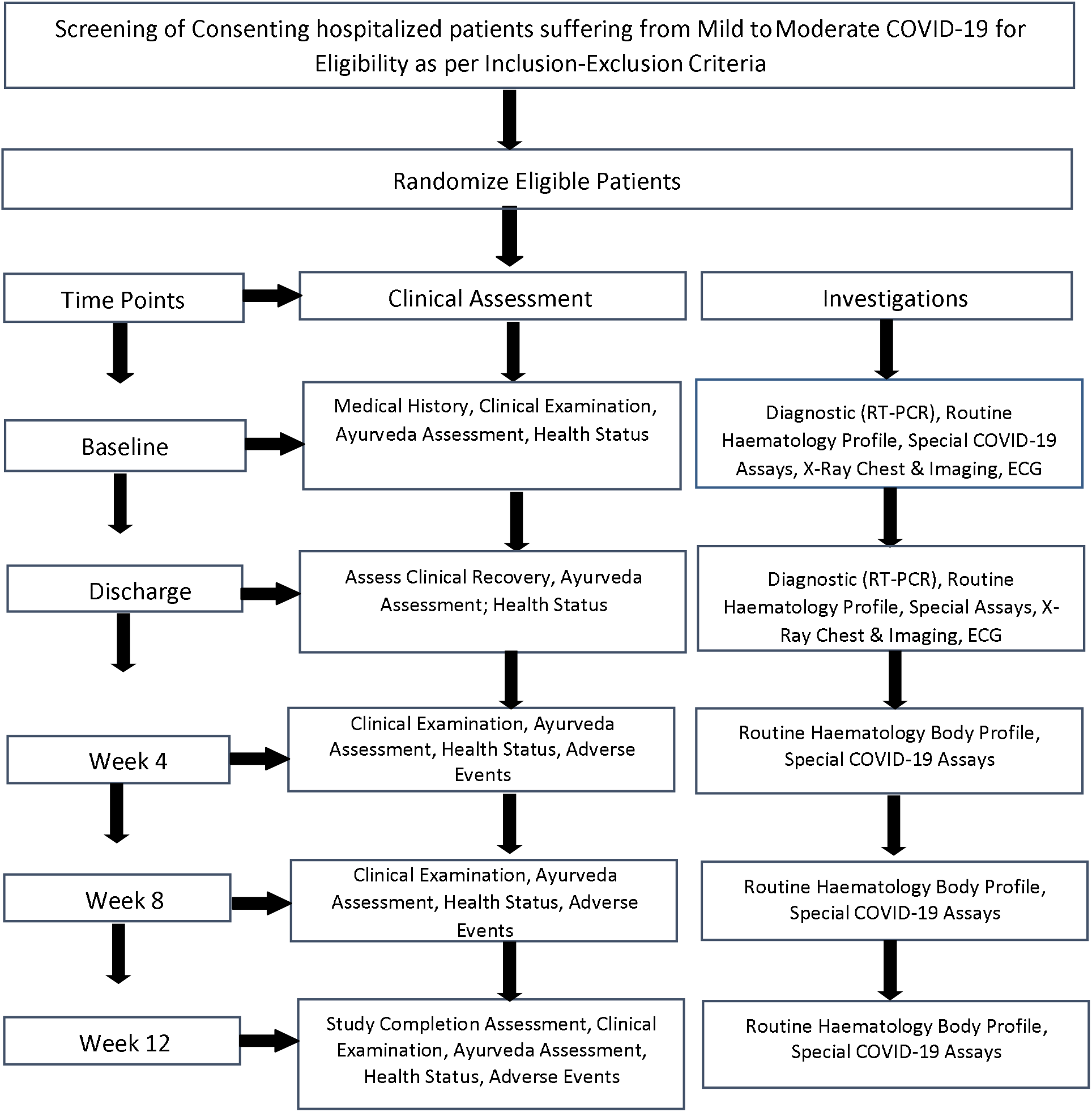
Study Flow diagram showing Study Events and Timelines: A randomized controlled study to evaluate the co-administration of AYUSH 64 with Standard of Care in COVID-19

### Patients

Voluntary hospitalized patients suffering from mild and moderate symptomatic COVID-19 were selected and after signing informed consent were screened for eligibility. Patients were jointly examined throughout the study by a modern medicine and Ayurveda physician (study investigator). All patients were managed for COVID-19 by an attending physician designated by the hospital at study site and who remained blinded to the treatment allocation in the trial (assessor blind). Patients and study personnel were aware of the treatment allocation (open label).

Adult patients with a typical clinical phenotype of COVID-19 illness and a laboratory confirmation test for SARS-CoV-2 (RT-PCR on a nasal and/or throat swab) were selected. Patients with severe symptomatic COVID-19 were excluded after fulfilling at least two criteria (i) respiratory distress at room ambience (ii) Oxygen saturation (SpO_2_) at rest ≤ 93% (iii) known COVID-19 complication. Full details of the inclusion and exclusion criteria can be accessed in the protocol registered at CTRI website (26).

### Trial Procedures

Patients at each site were randomized to a SOC arm or AYUSH 64 (investigational product) along with SOC (AYUSH plus) in 1:1 ratio on a first come first serve basis. A central randomization schedule was prepared by a biostatistician (SS) using a standard statistical software (WinPepi) and in blocks of 20 participant each. The randomized blocks were provided online with restricted access only to the principal investigator at each site as per the allotted sample size.

### Standard of care

All patients were begun on SOC soon after hospital admission as per the clinical judgement of the attending hospital physician. The concomitant use of hydroxychloroquine, azithromycin, corticosteroids, antibiotics, ivermectin, zinc, vitamin C, antiplatelet agents was as per the national guidelines of India but there were some local instructions as well (3). All patients were closely monitored on a daily basis as per standard COVID-19 protocol and which included respiration and peripheral arterial oxygen saturation, body temperature and blood pressure (3).

### Investigational drug

Each 500 mg tablet of AYUSH 64 contained aqueous extracts (100 mg each) of *Alstonia scholaris* (bark), *Picrorhiza kurroa* (rhizome), *Swertia chirata* (whole plant) and *Caesalpinia crista* (200 mg seed powder). The dose was two 500 mg tablets twice daily to be taken with a glass of water soon after a meal and this dosage remained fixed. Patients assigned to the AYUSH 64 arm continued the drug following clinical recovery till study completion (12 weeks). AYUSH 64 was procured from Indian Medicines Pharmaceutical Corporation Limited (IMPCL), Uttarakhand, India under arrangements with CCRAS, New Delhi. The manufacturing facility was certified ISO 9001 facility (2008) and followed guidelines of good manufacturing practice Ayurvedic Pharmacopoeia of India. Details of composition, quality standards and features of chemistry, manufacturing and controls are described in the <u>Supplementary File S1 (Table S1.1, Table S1.2, Table S1.3, Fig S1.1)</u>.

The assessment of general physical health, psychosocial health, and QOL was carried out by using the standard World Health Organization QOL BREF questionnaire (27) and a recently developed Health Related-Behaviour Habit and Fitness Questionnaire (HR-BHF CRD, Pune 2020 version).

The WHO QOL-BREF had 27 questions classified into 4 domains-physical health (Q 7-35), psychological health (Q 6-30), social relationships (Q 6-15) and environmental well-being (Q 8-40); range of score shown in parenthesis. Each question was answered on a 5-item categorical response; We used manually calculated raw scores (summation) for each domain and a higher score meant better health.

HR-BHF contains nine questions pertained to general health, anxiety, fatigue, energy level, bowel habits, stress, happiness, sleep and appetite (food). Each question was answered on a 100 mm horizontal visual analogue scale (VAS) which was anchored at either end (0 and 100 mm) for the worst or best outcomes. We used individual question scores (0-100) and total score (0-900) for analysis in this study.

A comprehensive description of WHO QOL BREF (Text Box S2.1) and HR-BHF questionnaire including pre-study validation (Text Box S2.2, Text box S 2.3) is provided in the <u>Supplementary File S2</u>.

Standard procedures as described by ICH-GCP and India guidelines were followed to classify, record and monitor, and assign causality to all adverse events (AE) (23,24). Safety was assessed in case of all patients who were randomized and included all those withdrawn for whatever reason. Other than clinical evaluations, routine laboratory measures (including haematocrit, metabolic hepatic and renal profile, urinalysis) were also carried out. Electrocardiography was recorded during screening, hospital discharge and study completion.

Skiagram chest was carried out on screening, discharge from hospital and study completion. Data was collected on a daily basis till discharge from the hospital following clinical recovery. Subsequently, the data was collected as per the predetermined follow up schedule at 4, 8 and 12 weeks shown in Fig 1. Data was recorded in study case report forms at point of care and later entered into a central electronic data base by designated study personnel under supervision of the site co-ordinator and an appointed CRO (contract research organization).

Patients were counselled about post-COVID care. A specially designed software program for mobile application called ‘COVID KAVACH’ was used to track the patients for any kind of symptom or untoward event on a daily basis after discharge from the hospital (28).

### Outcome measures

The primary efficacy measure was (i) the mean duration (days) from baseline randomization to day one of clinical recovery (CR) (ii) Proportion of patients showing clinical recovery within a time framework of 28 days. Clinical recovery was accepted when all of the following criteria were met with for least 48 hours under medical supervision of the attending physician (a) normal body temperature (≤36.6°C axilla or ≤37.2°C oral) (b) absence of any cough requiring any form of regular medication (c)absence of breathlessness on routine daily self-care activities and respiratory rate less than 30 breaths per minute without supplemental oxygen (d) absence of any other symptom/sign attributed to COVID-19 illness and requiring continuous treatment (e) normal SpO2 by standard peripheral oximetry device (above 94 percent)(f) negative RT-PCR assay for SARS-CoV-2 from nasal and throat swab. There were several secondary efficacy measures pertaining to (i) time lines such as mean duration from onset of symptoms to CR, mean duration from hospitalization to CR (ii) COVID-19 related blood assay biomarkers such as C-reactive protein (CRP), D-Dimer, Ferritin, interleukin-6.

### Study Withdrawals

Patients worsening clinically and requiring prolonged oxygen and/or intensive care was withdrawn from the study and continued routine management in the hospital. All patients who were withdrawn were thoroughly questioned and evaluated to determine the reason for withdrawal. However, patients were permitted to withdraw without assigning any reason.

### Statistical Analysis

A convenience study sample of 140 participants was finalized by AC and SS and considered adequate to address the study research questions. The data were analysed for central tendencies (mean, median), range, standard error, standard deviation and 95% confidence interval (95% CI). Data were tabulated and graphically shown using standard format and MS Excel. Statistical tests were carried out to compare treatment groups as per the distribution (normality) Student’s T test (normative), Mann-Whitney statistic (non-parametric), Chi-square statistic (categorical), ANOVA. 95% confidence interval (95% CI) of the difference between mean of the study arms was calculated for efficacy measures; significance at p < 0.05 (two sided). Both intent-to-treat and per protocol completer analysis were performed when appropriate. Standard statistical software programs were used (GraphPad InStat Version 3.6 and Confidence Interval Analysis software analysis, BMJ Group, London, 2003). The study arm of ‘AYUSH 64 plus SOC’ is referred to as ‘AYUSH plus’ and ‘SOC alone’ is referred to as ‘SOC’ in the current paper.

## RESULTS

Total 140 patients were randomized with 70 patients in each of the two study arms. Fig 2 shows the patient disposition and withdrawals.

**Figure 2:**
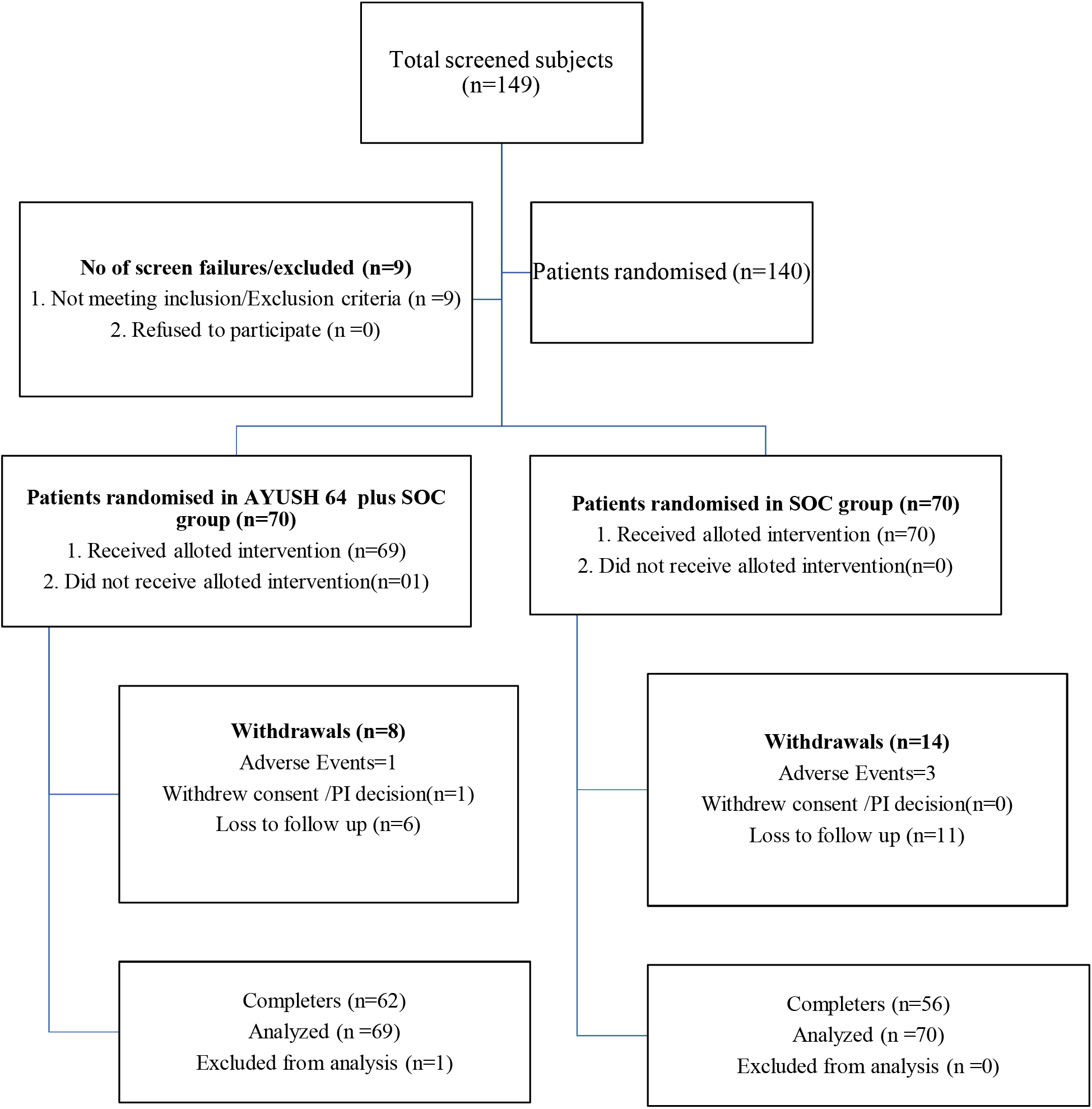
Patient disposition and withdrawals: a randomized controlled study to evaluate the co-administration of AYUSH-64 with Standard of Care (SOC) in mild - moderate symptomatic COVID-19 of 12 weeks duration (n=140)

One patient withdrew consent immediately after randomization. One patient in the SOC worsened and was withdrawn; recovered after intensive medical care. 138 patients completed treatment as per protocol. A total of 22 (15.7 %) patients were withdrawn (14 in SOC, 8 in AYUSH 64). 20 patients were withdrawn during the post recovery period; two patients with non-COVID related SAE (malaria and diabetes with severe cellulitis), 1 patient with acute onset mild polyneuropathy and later diagnosed as Guillain Barre syndrome, and 17 patients who refused to continue in the study following hospital discharge. None of the patients withdrawn reported a drug related AE. There were no deaths reported in the study.

### Randomization Baseline

Both the study groups were well matched for several demographic, clinical and laboratory variables as shown in Table 1. Most participants were men in the age range 30 -55 years. COVID-19 was classified mild in 80% participants. Common comorbid disorders were hypertension, known diabetes or first-time detected hyperglycaemia (fasting blood sugar > 120 mg/dl).

**Table 1:** Baseline data on demographic, clinical, selected laboratory variables and SOC drugs: A randomized controlled study to evaluate the co-administration of AYUSH 64 with Standard of Care in COVID-19

| Variables | AYUSH plus<br>(n = 69) | SOC<br>(n = 70) | Comparison,<br>p value* |
| --- | --- | --- | --- |
| **Clinical** |  |  |  |
| Age (years) Mean ± SD | 42.87 ± 12.6 | 42.7 ± 12.0 | 0.93 |
| Male – number (%) | 54 (77.14%) | 58 (82.85%) | 0.52 |
| Body Weight (kg) Mean ± SD | 69.34 ± 10.3 | 68.38 ± 12.1 | 0.61 |
| BMI ($\text{kg/m}^2$) Mean ± SD | 24.86 ± 3.4 | 24.53 ± 3.7 | 0.65 |
| Symptom onset to randomization (days), mean ± SD | 7.61 ± 4.8 | 7.83 ± 4.5 | 0.51 |
| Symptom onset to Hospitalization, mean ± SD | 6.4 ± 4.64 | 6.5 ± 4.47 | 0.75 |
| Hospitalization to Randomization, mean ± SD | 1.4 ± 0.8 | 1.5 ± 0.9 | 0.55 |
| Mild clinical disease-number (%) | 56 (80) | 58 (82.9) | 0.82 |
| Moderate clinical disease number (%) | 14 (20) | 12 (17.1) | 0.82 |
| Hypertension-number (%) | 17 (24.29) | 10 (14.29) | 0.19 |
| Diabetes mellitus-number (%) | 14 (20) | 06 (8.57) | 0.09 |
| Undiagnosed hyperglycaemia-number (%) | 9 (12.85) | 14 (20) | 0.36 |
| Blood sugar level mg/dl, mean ± SD | 112.50 ± 37.5 | 114.17 ± 35.2 | 0.74 |
| ESR mm fall 1<sup>st</sup> hour, mean ± SD | 50.2 ± 38.0 | 46.9 ± 37.4 | 0.79 |
| Blood haemoglobin gm/dl, mean ± SD | 13.6 ± 1.42 | 13.8 ± 1.62 | 0.51 |
| Total leucocyte count/cu mm, mean ± SD | 5920.7 ± 2008 | 6828.3 ± 2085 | 0.02 |
| Total Lymphocyte count/cu mm, mean ± SD | 32.31 ± 9.1 | 31.07 ± 09.6 | 0.34 |
| **Symptoms at baseline** |  |  |  |
| Fever | 53 (75.71%) | 45 (64.28%) | 0.10 |
| Sore throat | 46 (65.71%) | 53 (75.71%) | 0.24 |
| Cough | 54 (77.14%) | 54 (77.14%) | 0.87 |
| Dyspnea | 24 (34.28%) | 25 (35.71%) | 0.90 |
| Myalgia | 48 (68.57%) | 54 (77.14%) | 0.31 |
| Headache | 37 (52.85%) | 32 (45.71%) | 0.35 |
| Diarrhea | 11 (15.71%) | 12 (17.14%) | 0.85 |
| Ageusia | 19 (27.14%) | 19 (27.14%) | 0.96 |
| Anosmia | 13 (18.57%) | 14 (20%) | 0.86 |
| **Drugs administered\*\*** |  |  |  |
| Tab Azithromycin | 48 (70%) | 49 (70%) | 0.95 |
| Tab Doxycycline | 1(2%) | 0 | 0.31 |
| Tab HCQS | 29 (42%) | 24(34%) | 0.35 |
| Tab Zinc | 48(70%) | 42(60%) | 0.24 |
| Tab Vitamin C | 69 (100%) | 69(99%) | 0.32 |
| Tab Vitamin D 3 | 15(22%) | 18(26%) | 0.58 |
| Tab Pantoprazole | 66(96%) | 65(93%) | 0.73 |
| Tab Paracetamol | 59 (86%) | 55(79%) | 0.28 |
| Tab Cetirizine | 13(19%) | 15(21%) | 0.70 |
| Tab Ivermectin | 3 (4%) | 2 (2.9%) | 0.99 |
| Anti-coagulant | 13(18.8%) | 13(19%) | 0.97 |
| Oxygen*** | 9(13%) | 6 (9%) | 0.39 |
| Note: *p: significance p<0.05, two tailed (ANOVA, Chi Square statistic). #Others (less than 7 patients): easy tiredness, giddiness, nausea, vomiting, backache, abdominal discomfort; **All were standard tablets (Indian pharmacopeia) except anti-coagulant (injection low molecular weight heparin analogue) and intermittent oxygen (face mask); ***intermittent; n: number of study participants; SD: standard deviation; AYUSH plus: AYUSH 64 + SOC, Other comorbid disorders: hyperlipidaemia (2),cardiac disorder (1), chronic lung disease(2), thyroid disorder (5), Allergic rhinitis (3) ; BMI: body mass index; ESR: erythrocyte sedimentation rate (Wintrobe method);See text for detail |  |  |  |

The study arms were well matched for several timelines at randomization baseline such as’ onset of symptom to hospital admission’ (−1.34 to 1.72),’ hospital admission to randomization’ (−0.17 to 0.39) and ‘symptom onset to randomization’ (−1.08 to 1.98); 95% CI of the difference between the group means (days) shown in parenthesis (Table 1). Site specific data for selected timelines, including related to RT-PCR assay, is shown in <u>Supplementary File S3</u>, Table S3.2. Some of these timelines are likely to have influenced the primary efficacy measure (time period from randomization baseline to clinical recovery)

Importantly, the study arms were well matched for individual SOC drug intervention (Table 1); drug dosage (Table S 3.1) and site-specific data (Table S3.2) is shown in <u>Supplementary File S3</u>. Skiagram of chest was reported (by radiologist) normal in 47% patients, mild abnormalities in 51% patients and moderate abnormalities in 2% patients in the AYUSH plus; correspondingly 46%, 41% and 13% in the SOC.

### Efficacy

Four participants started AYUSH 64 after 36 hours of randomization and were excluded from the primary efficacy analysis; addition of their data did not materially change the outcome (data not shown).

The mean duration (days) for complete recovery (primary efficacy) from the randomization baseline was significantly superior in AYUSH 64 (6.5±2.4 days) as compared to SOC (8.3±4.4 days) and the 95% CI of the difference in means was -3.03 to -0.59 days (Table 2). The latter improvement was observed at each of the study site. A higher proportion of patients in the AYUSH 64 (69.75%) showed complete recovery as compared to SOC (52.9% patients) during the first week following randomization (p=0.046, Chi-square statistic).

**Table 2:**
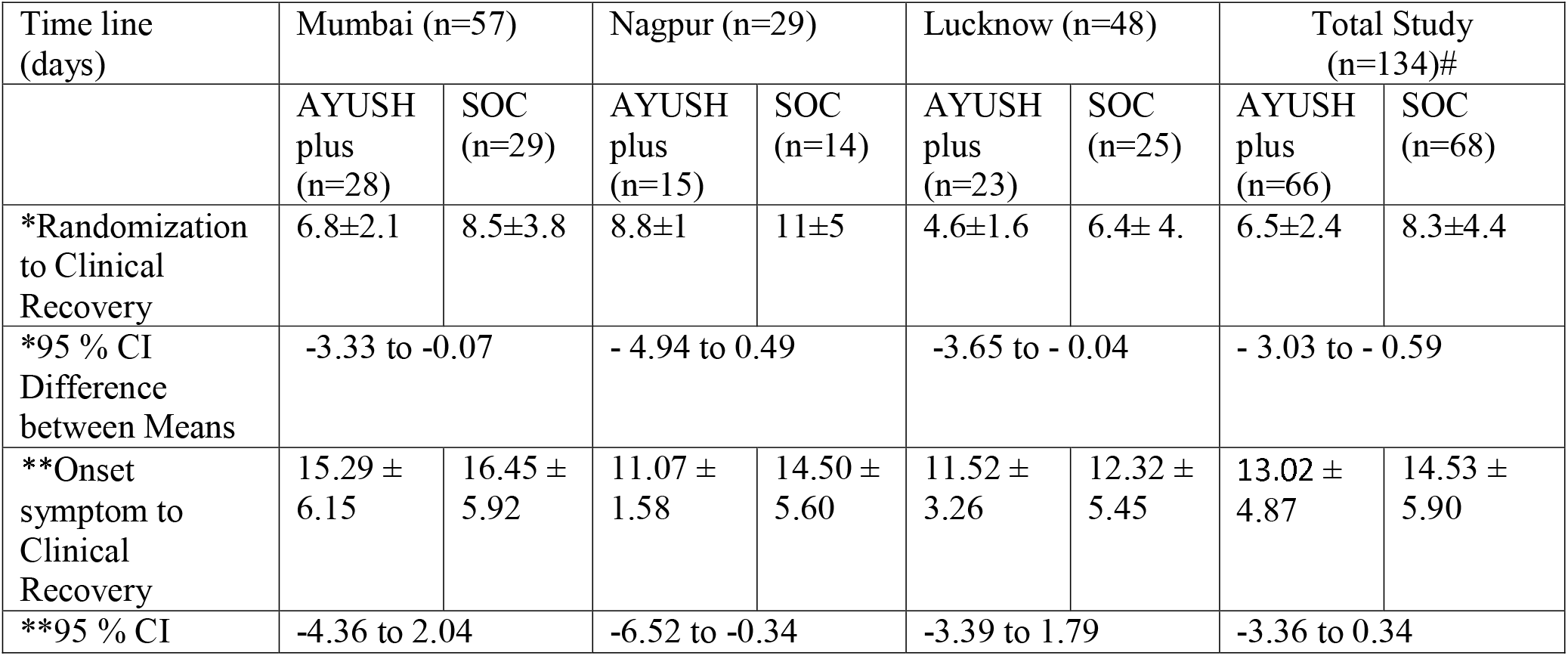
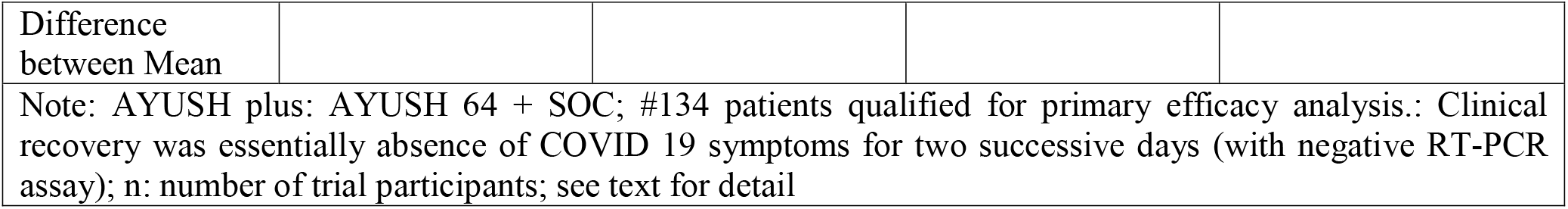
Primary efficacy measure (randomization to clinical recovery) and selected timeline: A randomized controlled study to evaluate the co-administration of AYUSH-64 with Standard of Care (SOC) in mild - moderate symptomatic COVID-19 (n=139);[Mean (days)± Standard Deviation]

| Time line (days) | Mumbai (n=57) |  | Nagpur (n=29) |  | Lucknow (n=48) |  | Total Study (n=134)# |  |
| --- | --- | --- | --- | --- | --- | --- | --- | --- |
|  | AYUSH plus (n=28) | SOC (n=29) | AYUSH plus (n=15) | SOC (n=14) | AYUSH plus (n=23) | SOC (n=25) | AYUSH plus (n=66) | SOC (n=68) |
| *Randomization to Clinical Recovery | 6.8±2.1 | 8.5±3.8 | 8.8±1 | 11±5 | 4.6±1.6 | 6.4± 4. | 6.5±2.4 | 8.3±4.4 |
| *95 % CI Difference between Means | -3.33 to -0.07 |  | - 4.94 to 0.49 |  | -3.65 to - 0.04 |  | - 3.03 to - 0.59 |  |
| **Onset symptom to Clinical Recovery | 15.29 ± 6.15 | 16.45 ± 5.92 | 11.07 ± 1.58 | 14.50 ± 5.60 | 11.52 ± 3.26 | 12.32 ± 5.45 | 13.02 ± 4.87 | 14.53 ± 5.90 |
| ***95 % CI | -4.36 to 2.04 |  | -6.52 to -0.34 |  | -3.39 to 1.79 |  | -3.36 to 0.34 |  |

|  |
| --- |
| Difference between Mean |
| Note: AYUSH plus: AYUSH 64 + SOC; #134 patients qualified for primary efficacy analysis.: Clinical recovery was essentially absence of COVID 19 symptoms for two successive days (with negative RT-PCR assay); n: number of trial participants; see text for detail |

The earlier recovery in the AYUSH 64 plus was also observed in case of ‘time to clinical recovery from onset of symptom’ but this was not significant compared to SOC (Table 2).

In both the study arms there was a significant reduction in COVID-19 serum biomarkers but there were no significant differences at randomization, clinical recovery and study completion (Table 3).

**Table 3:** COVID-19 biomarkers: a randomized controlled study to evaluate the co-administration of AYUSH-64 with Standard of Care (SOC) in mild - moderate symptomatic COVID-19 (n=139); [Mean (days)± Standard Deviation]

| Variable | Study groups | Baseline* | On discharge* | Week 12* |
| --- | --- | --- | --- | --- |
| Lactose dehydrogenase (LDH) | AYUSH plus | 403.6 ± 131.6 | 338.9 ± 109.7 | 318.7 ± 109.6 |
|  | SOC | 446.7 ± 206.5 | 363.8 ± 115.2 | 381.9 ± 164.8 |
| LDH Reference Range: 225-480 U/L |  |  |  |  |
| Ferritin | AYUSH plus | 337.8 ± 280.3 | 257.7 ± 226.1 | 84.4 ± 70.2 |
|  | SOC | 337.4 ± 278 | 201.8 ± 206 | 92.5 ± 89.3 |
| Ferritin Reference Range: Male 30-350 ng/ml; Female 20-250 ng/ml |  |  |  |  |
| Procalcitonin | AYUSH plus | 0.1 ± 0.1 | 0.1 ± 0.1 | 0.1 ± 0.2 |
|  | SOC | 0.1 ± 0.04 | 0.1 ± 0.2 | 0.1 ± 0.2 |
| Prolactin Reference Range: <0.2 ng/ml |  |  |  |  |
| C-reactive protein | AYUSH plus | 20.83 ± 27.55 | 10.3 ± 19.1 | 6.3 ± 6.5 |
|  | SOC | 25.5 ± 35.3 | 10.7 ± 12.5 | 6.39 ± 8.98 |
| C-Reactive Protein Reference Range <3 mg/L |  |  |  |  |
| D-Dimer | AYUSH plus | 462.5 ± 439.9 | 334 ± 224.9 | 297.3 ± 277.6 |
|  | SOC | 523.2 ± 672.8 | 345.3 ± 324.2 | 317.9 ± 418.4 |
| D-Dimer Reference Range:0-400 ng/ml |  |  |  |  |
| Interleukin-6 | AYUSH plus | 30.6 ± 46.0 | 7.7 ± 12.2 | 8.5 ± 22.1 |
|  | SOC | 32.6 ± 42.2 | 8.5 ± 15.8 | 7.4 ± 10.3 |
| Interleukin-6 Reference Range: Up to 7 pg/ml |  |  |  |  |
| Note: AYUSH plus: AYUSH 64 + SOC; n=number of participants; * : no difference between the groups at significant p <0.05 (Mann Whitney statistic); see text for detail |  |  |  |  |

At the time of clinical recovery/hospital discharge, skiagram of chest were reported normal in all except for mild abnormalities 22% patients AYUSH plus and 21% patients SOC; findings consistent with COVID-19. None of the patients with radiological abnormalities complained of fever, persistent cough or breathlessness during the post hospital discharge follow up. There were no clinical diagnosed post-COVID lung complications diagnosed on study completion; skiagrams for several patients were reported normal.

In comparison to SOC, AYUSH 64 plus showed significant improvement in several domains (physical health, psychological health, social relationship and environmental well-being) in the WHO QOL BREF and the total HR-BHF score on clinical recovery and during pre-determined follow up evaluations (Table 4). Of note was a significant superior improvement (Table S2.1) in AYUSH 64 in fatigue, stress, anxiety, appetite and happiness as compared to SOC using HR-BHF questionnaire and shown in <u>Supplementary File S2</u>.

**Table 4:**
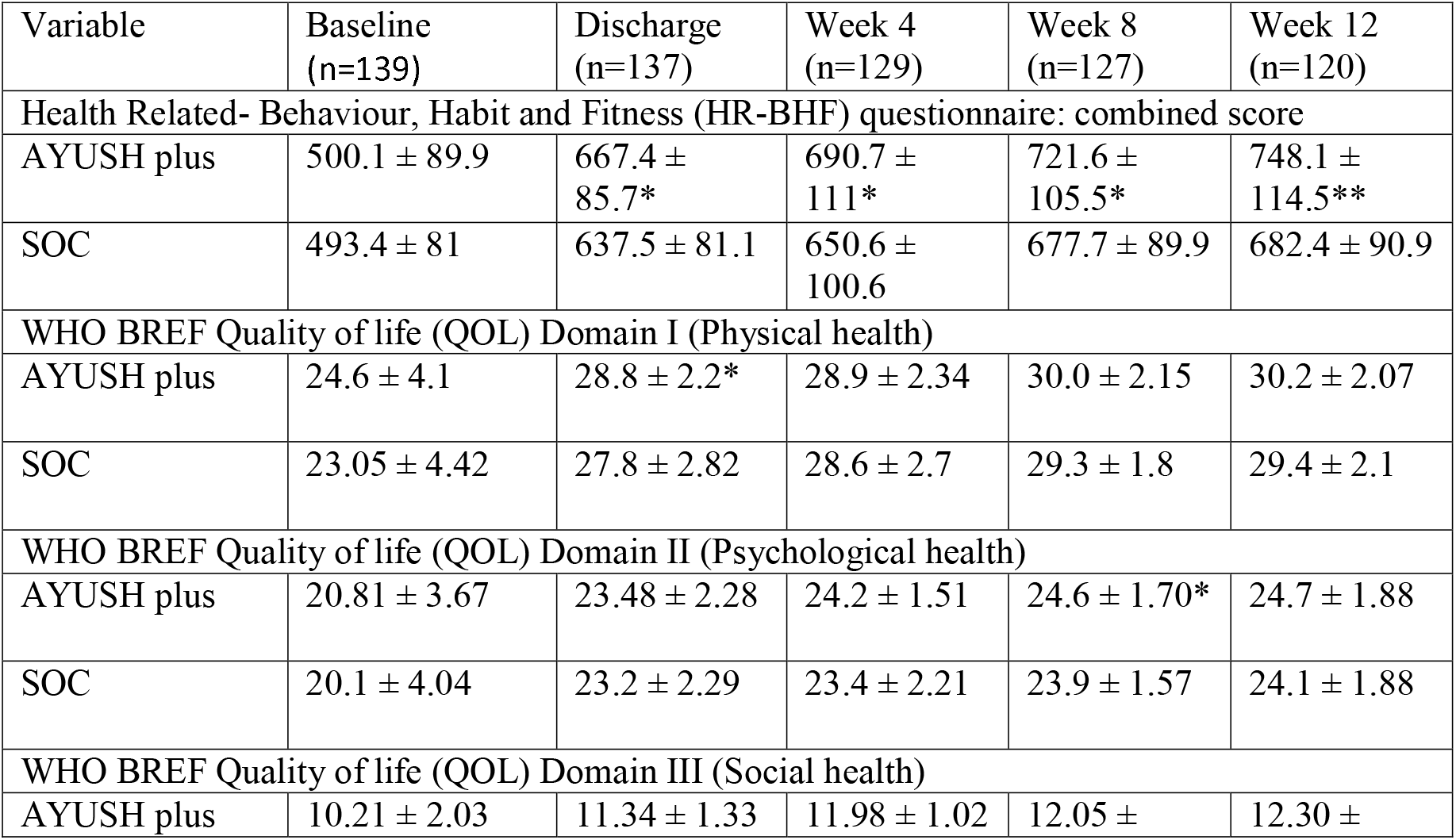
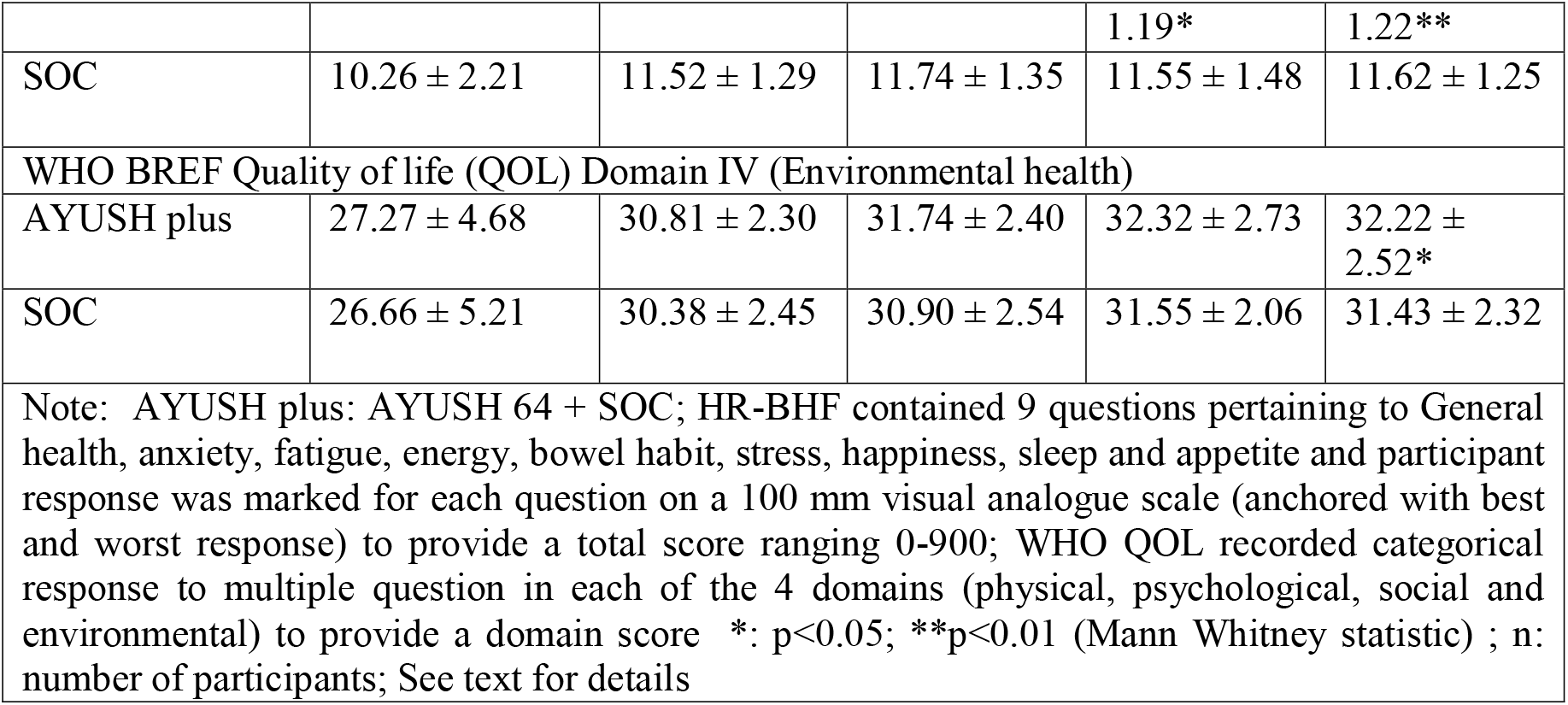
Quality of life questionnaires scores (Secondary outcome): a randomized controlled study to evaluate the co-administration of AYUSH-64 with Standard of Care (SOC) in mild and moderate symptomatic COVID-19 (n=139)

### Safety and related issues

A total of 49 AEs were recorded in 28 patients in the AYUSH plus and 52 AE in 29 patients in the SOC group: no significant differences (Table 5). Additional data on AE as per pre-determined study time points in each of the treatment arms is shown in <u>Supplementary File S4</u>, Table S4.1.

**Table 5:** Number of participants with adverse events (AE), causality and worsening in a randomized controlled study to evaluate the co-administration of AYUSH-64 with Standard of Care (SOC) in mild and moderate symptomatic COVID-19 (n=139)

| Details of Adverse Events | AYUSH plus (n=69) |  | SOC (n= 70) |  |
| --- | --- | --- | --- | --- |
|  | Participant | Events | Participant | Events |
| **Adverse events summary** |  |  |  |  |
| Fatal cases | 0 | 0 | 0 | 0 |
| Serious AEs | 0 | 0 | 3* | 3* |
| Adverse events | 29 | 48 | 26 | 48 |
| **Causal relation (WHO criteria of causation)** |  |  |  |  |
| Unrelated | - | 45 | - | ND |
| Unlikely | - | 03 | - | ND |
| **Adverse events (System organ classification)** |  |  |  |  |
| Cardiac | 1 | 1 | 0 | 0 |
| Ear and labyrinth | 0 | 0 | 1 | 1 |
| Gastrointestinal | 10 | 10 | 5 | 5 |
| Hepatobiliary | 0 | 0 | 1 | 1 |
| Infections and infestations | 11 | 12 | 10 | 11 |
| Skin and subcutaneous tissue | 2 | 2 | 1 | 1 |
| Respiratory, thoracic, mediastinal | 6 | 7 | 7 | 8 |
| Nervous system | 3 | 3 | 0 | 0 |
| Renal and urinary | 0 | 0 | 1 | 1 |
| Endocrine | 6 | 6 | 6 | 6 |
| Investigations | 0 | 0 | 1 | 1 |
| Others | 7 | 7 | 10 | 13 |
| Note: AYUSH plus: AYUSH 64+ SOC: n: number of participants; No AE recorded for disorders of blood and lymphatic, immune system, metabolism and nutrition, psychiatric, reproductive system and breast, eye, vascular system, congenital familial and genetic, injury poisoning and procedural complications, and surgical and medical procedures; See text for detail |  |  |  |  |

Several mild AE such as fever, myalgias, fatigue, breathlessness, loss of taste and or smell were considered related to COVID-19 and seems to persist in several participants at week 4 follow up. None of the AE were considered causally related to AYUSH 64 or any other drug used in the study. However, a higher number of gut related AE were reported in patients continuing AYUSH 64 after clinically recovery though definite causality could not be established. Only 3 patients in the SOC arm suffered from SAE which were not related to any drug. All other AE were considered mild and treated symptomatically; patients with naïve hyperglycaemia or dyslipidaemia were treated by specialist physicians. One patient in the SOC group developed mild peripheral polyneuritis following recovery which was later diagnosed Guillain Barre syndrome (GBS); patient recovered completely.

Repeated routine laboratory assays remained within normal limits in the two arms and there were no significant differences in the treatment arms (<u>Supplementary File S5</u>, Table S5.1). Electrocardiography was reported normal at baseline, hospital discharge and on study completion.

## DISCUSSION

This study compared a combination regimen of AYUSH 64 and SOC (AYUSH plus) with SOC to treat patients suffering from mild and moderate COVID-19 and admitted in the hospital.139 of the 140 randomized patients achieved complete recovery.118 patients completed 12 weeks of study duration. 69.7% patients in the AYUSH plus versus 51.7% patients in SOC arm recovered within the first week after randomization (p=0.046). The mean duration (days) to clinical recovery after randomization baseline in the AYUSH plus (6.5±2.4 days) was significantly superior to SOC (8.3±4.4); 95% CI of difference - 3.03 to - 0.59 days. Patients also showed better improvement for several secondary measures in AYUSH plus - reduced mean duration of clinical recovery from symptom onset and higher persistent scores of general physical and mental health and QOL. There were no significant differences in the AE by study arms. Except for three serious AE in the SOC group, all AE were mild and none were considered causally related to a study drug. 17 of the 22 patients withdrawn had refused to follow up after hospital discharge 1 patient withdrew consent soon after randomization and 1 patient in the SOC arm required critical medical care for recovery. There were no deaths in the study.

AYUSH 64 was continued till study completion and patients reported more gut related AE, albeit mild and with no definite causality. One patient is likely to have suffered from post COVID-19 complication of Guillain Barre Syndrome (29). None of the patients on follow up were diagnosed with pulmonary fibrosis. 24% of the study cohort had co-existent diabetes or naïve hyperglycaemia on randomization which often required adjustment in blood sugar medication (Table 1). All patients made uneventful recovery except for one patient of diabetes who recovered from COVID-19 but few days later developed severe cellulitis in the leg requiring extended hospitalization. Uncontrolled hyperglycaemia was reported to worsen the prognosis in COVID-19 (30).

### Study Implications

The national India policy regarding management of mild and moderate COVID-19 has shifted predominantly towards isolation in home or quarantine (31). The latter was meant to ‘free’ hospital beds for severe and critical cases. Several repurposed modern medicines (such as hydroxychloroquine and ivermectin) that were extensively used earlier in the pandemic are no longer favoured (4, 7). The oral treatment is largely empirical (1,2,3). The current AYUSH 64 drug trial needs to be viewed against this perspective.

The current study demonstrated early clinical recovery (COVID-19) when AYUSH 64 was combined with SOC. AYUSH 64 also showed good safety and persistent improvement in health over 12 weeks of use. Undoubtedly, AYUSH 64 can be recommended for use in domiciliary and quarantine setting but with a caveat. It needs to be medically supervised and patients suitably counselled as was done in the current study. It is difficult to predict severe COVID-19 (32). A small proportion of mild cases develop severe disease (32, 33). However, mild and moderate cases may develop post COVID-19 complications (34).

Provision of timely critical care in a hospital is a pivotal component for successful management of the pandemic. In our experience, despite sound medical advice, a large proportion of mild and moderate uncomplicated cases are admitted in the hospital and clog the system. AYUSH 64 plus SOC seemed to have significantly reduced the duration of hospitalization. A similar reduction in the length of hospital stay was reported in a meta-analysis of controlled drug trials of co-administration of Chinese herbal medicine with conventional western medicine in COVID-19 (95 % CI of the mean difference was -3.28 to -0.70 days) (8). COVID-19 is a dreadful disease with a huge burden of psychosocial disorders (35). A meta-analysis from India reported several psychological comorbidities ranging from 26% (anxiety and depression) to 40% (poor sleep quality) of study participants (36). The results of the current study were reassuring. Several patients reported early and persistent improvement that was superior in the AYUSH plus arm-reduced anxiety and stress, improved energy and general health and sleep, and more happiness (Table 4 and <u>Supplementary File S2</u>, Table S2.1). In our experience, patients found it easier to answer visual analogue scale-based questions in the HR-BHF dealing with mental and QOL issues as compared to the somewhat cumbersome standard WHO QOL BREF instrument.

The current study lends credence to the therapeutic potential of AYUSH 64. The plant ingredients are mentioned in ancient Ayurveda texts and have been traditionally used to treat a wide variety of illnesses and medical disorders in several Southeast Asian countries, China, Europe and North America (37, 38, 39, 40, 41). Several experimental biological effects described for these plants seem to be of clinical relevance to the present study (42, 43, 44). Recently, an impressive inhibition of a key replication protease enzyme in SARS-CoV-2 by AYUSH 64 ingredient was demonstrated in an in-silico molecular docking study (45).

### Strengths and Limitations

The current study repurposed a standardized Ayurvedic polyherbal formulation in popular Ayurveda (India) practise for over three decades to improve the efficacy of SOC treatment in COVID-19. The plant ingredients of AYUSH 64 have been in traditional ancient medicinal use in India and several countries.

In the current study, patients were directly observed on a daily basis in a hospital setting leading to good compliance and robust data capture. Despite serious concerns, the study arms were well matched on several measures including SOC. The results of primary outcome and safety were consistently in favour of AYUSH plus at each study site. AE were generally mild and not shown to be definitely caused by AYUSH 64. Importantly, patients showed persistent improvement in physical and mental health and quality of life. None of the patients in AYUSH plus arm worsened and there were no deaths in the study cohort We believe that the current study has boldly addressed the need for evidence-based medicine to treat mild and moderate COVID-19.

Several limitations were imposed by the chaotic and tragic pandemic situation. We encountered uncertainties and often contradictory advice regarding SOC and other COVID-19 related health matters in the social and news media. During the first pandemic year, the patients were reluctant to seek medical care for fear of being stigmatized and this probably delayed the treatment for several patients as shown by the timelines in Table 1. Though, the primary efficacy was assessed by the attending physician in a blinded manner, a placebo response to some extent cannot be ruled out. Ayurveda is endearing to the Indian community. In view of absence of a-priori data, we settled for a convenience sample size for this study.

### Other Studies

Several drug trials using Ayurveda and other CAM therapies in COVID-19 were registered in the clinical trial registry of India but a few are published (36, 46). A large effort has been put in by the Ministry of AYUSH, GOI, to validate these drugs (21). A randomized placebo controlled short term study attempted to show a reduction in viral load using a well-designed Ayurvedic regimen (oral drugs and nasal application) in asymptomatic and mild cases of COVID-19; clinical improvement was not properly defined (47). A small sample size uncontrolled observational study showed good resolution of symptoms by AYUSH 64 in patients suffering from influenza like illness (48).

Recently a cocktail use of monoclonal antibodies (MAB) have been demonstrated in controlled drug trials to show a substantial reduction in the viral load and clinical improvement in patients suffering from early COVID-19 and none of the study participants progressed to severe disease (49,50). The latter studies required MAB intervention within 24-48 hours of the illness onset and a firm diagnosis. This and several other restrictions described in the drug trial studies are likely to complicate widespread acceptance and clinical use, and more so in settings such as ours with several limitation in socioeconomics and medical care (49,50). Oral drugs like AYUSH 64 are a much more attractive proposition.

The recent data on the use of repurposed and adjuvant drugs in hospital patients with COVID-19 did not make any mention of herbal drugs or other CAM therapies (51). It is likely that the use of Ayurveda and other CAM therapies such as AYUSH 64, both in the hospital and community setting, was several folds more and we need more suitable data (18).

### Future Research

AYUSH 64 needs to be evaluated in a suitable designed phase III drug trial. The latter study should also evaluate the likely role of AYUSH 64 to block progression to severe disease and reduce post COVID-19 complications. Experimental evidence is required to validate its anti-viral and salubrious effect on health and QOL.

## CONCLUSION

In a randomized controlled prospective study of hospitalized patients suffering from mild and moderate COVID-19, AYUSH 64 (a standardized polyherbal Ayurveda drug) was shown to be a significantly effective and safe adjunct to SOC. It hastened clinical recovery, reduced hospitalization period and showed early persistent substantial benefit towards physical and mental health.

## Data Availability

Not applicable

http://ctri.nic.in/Clinicaltrials/showallp.php?mid1=43812&EncHid=&userName=025557

## Authors Contribution

The protocol was prepared by AC, GT and MS. The interpretation of data was principally carried out by AC in discussion with GT, MS, SS and BP (senior author) while writing the first draft of the manuscript. All the lead study investigators (MG, KC, GR, AS, ML, MG, HR) confirmed local site data and observations and data used in this report. The sections on the standardization, quality control and traditional medicinal use of AYUSH 64 were written by GT, NS, BCS, BJ, AR. The biostatistical analytical data and its verification in several instances was carried out by SS and ST. The authors had access to the study data and other observations and tables in the current report and vouch for the veracity of the data and the study report. The current manuscript was approved by all the authors. All the authors provided consent to submit this manuscript.

## Transparency statement

AC (lead author) affirms that the manuscript is an honest, accurate, and transparent account of the study being reported and that no important aspects of the study have been omitted. There were no discrepancies from the study as originally planned and documented in the protocol.

## Procedural and Participation Matters

The sponsor did not participate in the preparation of the study protocol. The sponsor had no role in collection of data, setting up study e-database, statistical analysis and interpretation of the results and the decision to submit the article for publication. The sponsor (CCRAS) is the proprietor of AYUSH 64 which is the investigational product in the current study. Several plant related details of standardization, quality control and chemistry manufacturing control (CMC) of AYUSH 64 were obtained from the sponsor.

A contract research organization (CRO) was appointed by the sponsor as per the standard operating procedure of the Government of India. The CRO was responsible for the general administration and logistics of the study, ensure site training and GCP compliance, collect data and enter into a central electronic database The CRO performed the data lock prior to data analysis. The first run of statistical analysis was done by an independent biostatistician (VP) appointed by the CRO. Pivotal statistical analysis was rechecked by SS, an independent senior biostatistician.

Amongst the authors, BCSR, BJ and NS were senior Ayurvedic physicians and research executives with CCRAS, and KC, HR, AS and MG were Ayurvedic physician scientists working in Ayurvedic institutions administered by CCRAS. All other authors (AC, GT, MS, AR, ST and BP) were independent professionals belonging to Ayurveda or modern medicine or modern science and had no contractual relationship with CCRAS or its institutions.

## Acknowledgement

A special thanks to Vaidya Dr Rajesh Kotecha, Secretary, Ministry of AYUSH, Government of India, for his invaluable guidance and inspiration towards completion of the current study project and preparation of study publications. We are grateful to senior Vaidya KS Dhiman, former DG CCRAS, GOI for speedy completion of drug trial. We thank several research colleagues in the CCRAS - Dr Ravindra Singh, Dr Shruti Khunduri and Dr B S Sharma. We acknowledge with gratitude several colleagues from each trial site- Dr Manish Deshmukh, Dr Swati Munde, Dr Pratap Makhija, Dr Alia Rizvi, Prof Wahid Ali, Dr. Neeta Warty, Dr. Parth Dave and Mr Akash Saggam. We make a special mention of Dr Manesha Talekar, Ayurvedic physician, who volunteered to be a co-investigator despite pregnancy and contracted COVID-19 during her medical duty. Dr Vinay Pawar played an important role in statistical analysis. Finally, we thank all the patients with folded hands for their participation and wholehearted support.

