## Supplementary File for "Coadministration of AYUSH 64 as an adjunct to Standard of Care in mild and moderate COVID-19: A randomised, controlled, multicentric clinical trial"

**Supplementary File S1: Composition, Chemistry, Manufacturing and Controls of AYUSH-64**

**Table S1.1: Composition of AYUSH-64 (standard proprietary Ayurvedic drug); contents of a 500 mg tablet**

| **S.No.** | **Name of the ingredient** | **Botanical name** | **Part used** | **Quantity** |
| --- | --- | --- | --- | --- |
| 1 | Saptaparna (Aqueous extract) | *Alstonia scholaris* | Bark | 100 mg |
| 2 | Kutaki (Aqueous extract) | *Picrorhiza kurroa* | Rhizome | 100 mg |
| 3 | Kiratatikta (Aqueous extract) | *Swertia chirata* | Whole plant | 100 mg |
| 4 | Latakaranja powder | *Caesalpinia crista* | Seed | 200 mg |

**Table S1.2:** Quality Standard (Specifications) of AYUSH-64 (standard proprietary Ayurvedic drug) and its ingredients for QC Analysis of AYUSH-64 and its ingredients

| **Sr** | **Test Parameters** | **Ingredients** | | | | **Formulation** |
| --- | --- | --- | --- | --- | --- | --- |
|  |  | ***Saptaparna* (Aqueous Extract)** | ***Kutaki***  **(Aqueous Extract)** | ***Chiraita***  **(Aqueous Extract)** | ***Latakaranja* (Seed Powder)** |  |
| 1 | Loss on drying | Not more than 9% | Not more than 6% | Not more than 8% | - | Not more than 6% |
| 2 | pH (1% Sol) | 4.5-6.5 | 4.0-7.0 | 5.0-7.0 | - | 4.0-6.5 |
| 3 | Total Ash | Not more than 12% | Not more than 5% | Not more than 15% | Not more than 5% | Not more than 25.0% |
| 4 | Acid insoluble Ash | Not more than 2% | Not more than 1% | Not more than 2% | Not more than 1% | Not more than 8.0% |
| 5 | Alcohol soluble extractive | Not less than 3% | Not less than 3% | Not less than 12% | Not less than 26% | Not less than 5.0 % |
| 6 | Water Soluble extractive | Not less than 85% | Not less than 80% | Not less than 80% | Not less than 4% | Not less than 30.0% |
| 7 | Heavy Metals  (Max. limit) | Lead 10 ppm  Arsenic 3.0 ppm  Mercury 1.0 ppm  Cadmium 0.3 ppm | | | | |
| 8 | Microbial Count | Total Microbial plate count = < 10^5^cfu/gm  Yeast and mould count = <10^3^cfu/gm | | | | |
| 9 | Specific Pathogens | *Escherichia* coli.**-** absent  *Salmonella* spp. *-* absent  *Staphylococcus aureus -* absent  *Pseudomonas -* absent | | | | |
| 10 | Aflatoxins | B1 = Not more than 0.5%  B2= Not more than 0.1 %  G1= Not more than 0.5%  G2= Not more than 0.1 % | | | | |
| 11 | Pesticide Residue | As per Annexure-I | | | | |

Note- Quality control and safety parameters of the ingredients and the formulation are complied with Ayurvedic Pharmacopoeia of India (API) limits/ In-house limits.

Fig S1.1 : HPTLC Chemo Profiling and UV spectrophotometric analysis of Ayush-64 (standard proprietary Ayurvedic drug) & its raw ingredients.

| 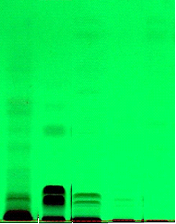 | 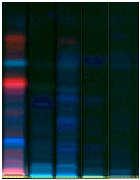 | 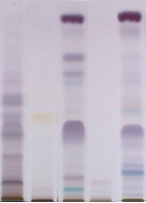 |
| --- | --- | --- |
| 1 2 3 4 5 1 2 3 4 5 1 2 3 4 5  UV 254 UV 366 under white Light after derivatization | | |

Track 1- Saptaparna , Track 2-Katuki , Track 3-Ayush 64, Track 4-Kiratatikta, Track 5-Latakaranja

(b)

| 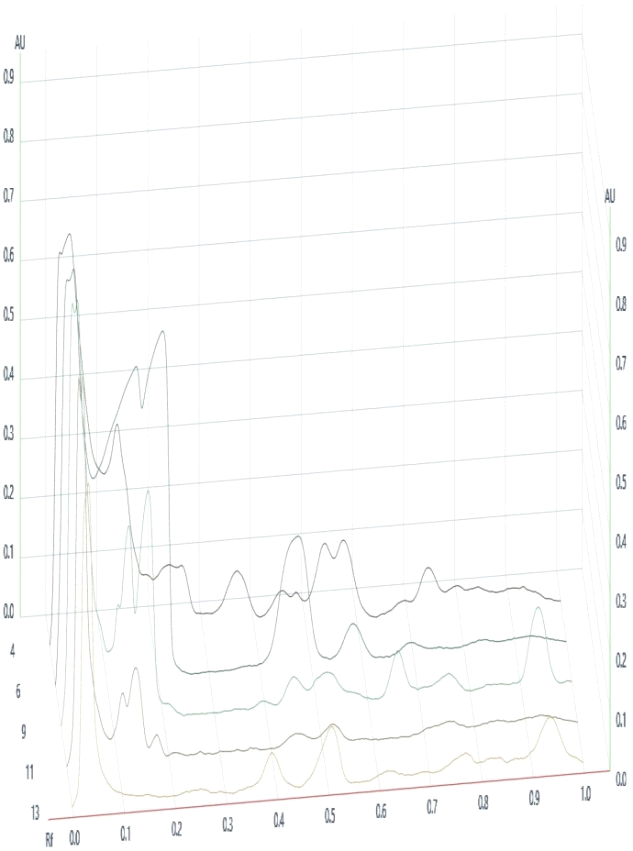 | 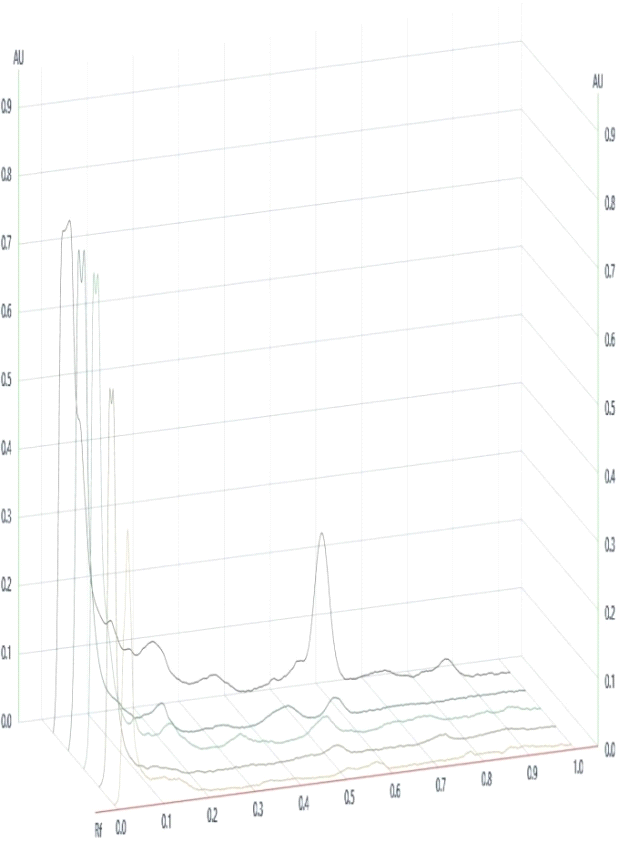 |
| --- | --- |

**3D Densiometric HPTLC Profile under UV 254 nm. 3D Densiometric HPTLC Profile under UV 366 nm.**

**Table S1.3:** Pesticide Residue of ingredients of AYUSH 64 (standard proprietary Ayurvedic drug) as per Indian pharmacopeia

| **Substance (s)** | **Permissible Limit prescribed in API** (in ppm) |
| --- | --- |
| Alachlor | 0.02 |
| Aldrin and Dieldrin (sum of) | 0.05 |
| Azinphos-methyl | 1.0 |
| Bromopropylate | 3.0 |
| Chlordane (sum of cis-,Trans- and Oxythlordane) | 0.05 |
| Chlorfenvinphos | 0.5 |
| Chlorpyrifos | 0.2 |
| Chlorpyrifos-methyl | 0.1 |
| Cypermenthrin (and Isomers) | 1.0 |
| DDT (sum of p, p-DDT, o, p-DDT, p, p-DDE and p, p-TDE) | 1.0 |
| Deltamenthrin | 0.5 |
| Diazinon | 0.5 |
| Dichlorvos | 1.0 |
| Dithiocarbamates (as CS2) | 2.0 |
| Endosulfan (sum of isomers and endosulfansulphate) | 3.0 |
| Endrin | 0.05 |
| Ethion | 2.0 |
| Fenitrothion | 0.5 |
| Fenvalerate | 1.5 |
| Fonofos | 0.05 |
| Heptachlor (sum of Heptachlor and Heptachlorepoxide) | 0.05 |
| Hexachlorbenzene | 0.1 |
| Hexachlorocyclohexane isomers (other than ɤ) | 0.3 |
| Lindane (ɤ- Hexachlorocyclohexane) | 0.6 |
| Malathion | 1.0 |
| Methidathion | 0.2 |
| Parathion | 0.5 |
| Parathion-methyl | 0.2 |
| Permethrin | 1.0 |
| Phosalone | 0.1 |
| PiperonylButoxide | 3.0 |
| Pirimiphos-methyl | 4.0 |
| Pyrethrins (sum of) | 3.0 |
| Quintozene (sum of pentachloroaniline and methyl pentachlorophnylsulphide) | 1.0 |

^$^ Limit of Detection = 10 ppb (GC-MS MS)

**Supplementary File S2: Quality of Life Questionnaires**

**Text Box S2. 1: A summary of the WHO Quality of Life (QOL)-Bref Questionnaire used in the randomized controlled study to evaluate the co-administration of AYUSH 64 (Ayurvedic drug) with standard of care in mild - moderate patients of COVID-19.**

WHO QOL-Bref (Dec 1996 version) has 27 questions and each question was answered on a 5 item categorical response scale (‘nil problem’ to ‘optimum problem’, score range 1-5) (22). The questions are classified into 4 domains- physical health (such as activities of daily living, sleep, work capacity), psychological health ( such as bodily image, positive feelings, negative feelings, self- esteem, spirituality), social relationships (such as personal relations, social support, sexual activity) and environmental wellbeing (such as finances, physical safety, health and social care, home environment, transport).

The total score for each domain varied (physical 7-35, psychological 6-30, social 6-15 and environmental 8-40).

Suitable local translations of the questionnaire in Hindi and Marathi language were used by the patients to record their response in the current study under reference.

In this study, we used manually calculated raw scores (summation) for each domain and a higher score meant better health. Reference: WHOQOL-BREF : Introduction, Administration, Scoring And Generic Version Of The Assessment. Field Trial Version, December 1996. Available at https://www.who.int/mental_health/media/en/76.pdf. Accessed on 26 May 2021

**Text Box S2.2: Health Related-Behaviour, Habit and Fitness Questionnaire (HR-BHF, CRD, Pune 2020 version)**

CONFIDENTIAL

ARTHRTIS RESEARCH CARE FOUNDATION-CENTRE FOR RHEUMATIC DISEASES PUNE INDIA HEALTH RELATED-BEHAVIOUR HABIT FITNESS (HR-BHF)

PARTICIPANT NO:	PARTICIPANT ID	Age	Gender

PARTCIPANT NAME (OPTIONAL)	Mobile (optional)	Address (General location, include name of village/city & district)

DATE:

Note: The participant is permitted to stay anonymous and not provide any contact details- only ID will suffice.

This is a research study. It will be used in clinical practice after testing its usefulness and acceptance by community and doctors.

Please put a vertical line across the scale to indicate your extent of the health-related problem

(i) **Please consider YOUR EXPERIENCE OVER THE PAST ONE WEEK while answering. We want you to give your overall assessment.**
1) **GENERAL HEALTH: How would you rate your overall general health? 0	100**

Very Poor	Very Good

2) **How much is your anxiety?**

0	_100

Very anxious	No anxiety at all

3) **Fatigue: How much do you feel tired in your daily routine?**

0	_100

Nil	Feel very tired

4) **How much is your Energy level?**

0	_100

Low energy	Feel Energetic during my daily routine

5) **How is your Bowel movement (passing stools)?**

0	100

Highly irregular and unsatisfying	Satisfactory and normal bowel habit

6) **Are you stressed?**

0	_100

No Stress	Maximum Stress

7) **How happy are you?**

0	100

Very Sad	Very Happy

8) **How do you rate your sleep?**

0	100

Normal	Bad and unable to sleep properly

9) **How much is your appetite for food?**

0	100

Nil	Very Good

REMARKS

### THANK YOU FOR YOUR PARTICIPATION. You may contact us at +912026344099/26345624 or

***and visit us at*** [***www.rheumatologyindia.org***](http://www.rheumatologyindia.org/)

Note that the above illustration is not drawn to scale.

**Text Box S2. 3: Development and Validation of Health Related-Behaviour, Habit and Fitness Questionnaire (HR-BHF CRD Pune 2020 version)**

HR-BHF was developed by Centre for Rheumatic Disease CRD), Pune in Feb 2020 to address the various complaints of general health related physical and mental health issues which are often associated with chronic disorders and often ignored by the doctors. But patients consider them important and they effect outcome. The latter was recognized by the rheumatologist (AC) in CRD. A new quality of life (QOL) assessment instrument namely HR-BHF (CRD Pune 2020) was developed. Inadvertently, this exercise coincided with the COVID 19 pandemic. We also speculated HR-BHF to be useful in assessing recovery in COVID 19. A summary along with the instrument is presented (results not yet published).

An inventory of 23 questions was created based on the information gathered from patients suffering from chronic arthritis and attending CRD outpatient from Dec 2019 to Feb 2020. A local CRD expert group of 3 physicians, 4 paramedics, 10 healthy community (5 senior citizens) and 12 patients decided to reduce the number of questions 9 after deliberate discussions and consensus. There were 14 women members in the expert group. The group opined that the questionnaire is likely to capture some vital aspects of human behaviour, habits and fitness (more mental than physical) and improve patient satisfaction. The face validity and suitable translations (local language) were confirmed by this expert group.

The 9 questions pertained to general health, anxiety, fatigue, energy level, bowel habits, stress, happiness, sleep and appetite (food). Patients marked the answer on a visual analogue scale (VAS) to indicate a measure of the difficulty or health status. VAS was a 100 mm horizontal bland line (no tick marks) anchored at 0 and 100 mm at either end for the extreme outcome. The patient drew a a short vertical mark on the VAS to record response and the intersection point was accurately measured from ‘0’ to provide the score. Addition of individual scores led to a composite HR-BHF score (range 0-900). The optimum response (best of health) for 5 questions (general health, energy happiness, bowel habit and appetite) was 100 and for 3 questions (anxiety, stress, depression) it was ‘0’ and this was decided after due consideration of the way patients understand the question.

Subsequently, an in-house evaluation of performance of HR-BHF was performed by AC and colleagues. 403 consenting participants [103 patients of chronic rheumatoid arthritis (RA) and 300 healthy community (HC] completed the questionnaire in face to face interview with a paramedic. 72.8% RA and 88.3% HC did not report ‘any difficulty’ in answering the questions; remaining had ‘some difficulty’. 100% RA and 98.4% HC reported HR-BHF to be ‘Useful to very useful’. The mean VAS score was 56.4 mm for general health, 43.6 mm for anxiety, 45.9 mm for fatigue, 55.2 mm for energy, 67.6 mm for good bowel clearance, 34.1 mm for stress, 67.9 mm for happiness, 38.8 mm for sleep and 68.7 mm for appetite in the RA group; correspondingly it was 81.7 mm, 34.7 mm, 27.9 mm, 77.1 mm, 76.7mm, 20.1 mm, 79.4mm, 33.5 mm, and 81.8 mm in the HC and significantly different (p<0.05, Student t test) for all measures except sleep. In the correlation matrix, general health was shown positively correlated with energy, good bowel clearance, happiness and good appetite and negatively correlated with anxiety, fatigue, stress and sleep. The correlation between each of the item questions varied from -0.126 to 0.503. 8 questions (HR-BHF) could explain 32.7% (adjusted R^2^ ) variation in the score of ‘general health’ (dependent variable, 9^th^ question in HR-BHF) in a multivariable regression model; data from the total cohort of 403 participants was used. The evaluation results were consistent with the content validity and clinical usefulness.

HR-BHF was used in the current drug trial of AYUSH 64 (Ayurvedic drug) and standard of care in mild and moderate COVID-19 as per protocol. The score at several time points was analysed to show response to intervention and recovery. The results of HR-BHF were also consistent with the standard WHO-QOL Bref questionnaire (See main text Table 8 and supplement material Table 5) .

The HR-BHF questionnaire is shown in supplement material Box 2

.

**Table S2.1: Individual question score (mean ± standard deviation) in Health Related- Behaviour, Habit and Fitness (HR-BHF, CRD Pune 2020 version) questionnaire: a** **randomized controlled study to evaluate the co-administration of AYUSH-64 with Standard of Care (SOC) in mild - moderate symptomatic COVID-19 (n=139)**

| Variable | Baseline (n=139) | | Discharge  (n=137) | Week 4  (n=129) | Week 8  (n=127) | Week 12  (n=120) |
| --- | --- | --- | --- | --- | --- | --- |
| HR-BHF-General Health | | | | | | |
| AYUSH plus | 60.85 ± 13.5 | 76.69 ± 10.0 | | 81.27 ± 9.8 | 84.87 ± 8.9 | 89.31 ± 8.6 |
| SOC | 58.29 ± 17.5 | 73.88 ± 13.3 | | 79.97 ± 12.0 | 83.57 ± 10.4 | 86.53 ± 9.2 |
| HR-BHF -Anxiety | | | | | | |
| AYUSH plus | 46.05 ± 21.1 | 67.02 ± 23.1 | | 61.75 ± 32.0 | 65.19 ± 33.7 | 68.74 ± 34.8* |
| SOC | 43.48 ± 22.4 | 69.22 ± 16.9 | | 58.95 ± 32.3 | 61.59 ± 33.1 | 57.79 ± 36.1 |
| HR-BHF-Fatigue | | | | | | |
| AYUSH plus | 48.06 ± 19.5 | 24.60 ± 14.0* | | 26.89 ± 17.1 | 21.02 ± 19.1 | 15.08 ± 13.4* |
| SOC | 51.91 ± 18.7 | 32.5 ± 18.5 | | 28.07 ± 18.7 | 22.09 ± 18.2 | 21.69 ± 19.4 |
| HR-BHF-Energy | | | | | | |
| AYUSH plus | 54.34 ± 18.2 | 72.26 ± 13.7 | | 77.63 ± 14.9 | 82.23 ± 13.1 | 87.65 ± 10.8 |
| SOC | 57.5 ± 17.0 | 71.98 ± 9.6 | | 77.55 ± 13.4 | 79.93 ± 14.2 | 83.62 ±15.2 |
| HR-BHF-Bowel movement | | | | | | |
| AYUSH plus | 60.97 ± 19.6 | 74.44 ± 12.0 | | 79.68 ± 16.1 | 83.94 ± 10.0 | 85.11 ±13.8 |
| SOC | 64.6 ± 19.2 | 76.38 ± 10.8 | | 78.34 ±15.6 | 80.66 ± 14.7 | 82.74 ± 12.1 |
| HR-BHF-Stress | | | | | | |
| AYUSH plus | 44.15 ± 20.6 | 21.13 ± 14.0* | | 19.81 ± 15.4 | 17.37 ± 15.1 | 15.69 ± 16.6 |
| SOC | 45.69 ± 21.1 | 26.47 ± 16.2 | | 22.29 ± 17.5 | 21.52 ± 17.6 | 18.09 ± 17.1 |
| HR-BHF-Happiness) | | | | | | |
| AYUSH plus | 56.29 ± 21.7 | 79.44 ± 15.1** | | 83.92 ± 12.2** | 86.02 ± 10.3** | 88.58 ± 8.1** |
| SOC | 57.84 ± 20.3 | 60.52 ± 31.2 | | 58.59 ± 31.2 | 61.52 ± 35.6 | 61.28 ± 36.9 |
| HR-BHF-Sleep | | | | | | |
| AYUSH plus | 48.89 ± 20.5 | | 32.82 ± 21.8 | 29.73 ± 28.0 | 29.16 ± 28.0 | 28.16 ± 31.3 |
| SOC | 54.36 ± 22.0 | | 30.17 ± 18.5 | 33.1 ± 30.4 | 29.5 ± 26.8 | 32.91 ± 31.4 |
| HR-BHF questionnaire -Appetite | | | | | | |
| AYUSH plus | 62.69 ± 17.2 | | 76.13 ± 12.5 | 83.06 ± 13.3 | 85.95 ± 8.0 | 87.65 ± 8.3* |
| SOC | 63.88 ± 18.5 | | 74.31 ± 10.5 | 80.67 ± 14.9 | 83.57 ±11.2 | 83.48 ± 12.3 |
| Note: AYUSH plus: AYUSH 64 + SOC; HR-BHF contained 9 questions pertaining to General health, anxiety, fatigue, energy, bowel habit, stress, happiness, sleep and appetite and participant response was marked for each question on a 100 mm visual analogue scale (anchored with best and worst response) to provide a total score ranging 0-900; *: p<0.05; **p<0.01 (Mann Whitney statistic) ; n: number of participants; Low scores for fatigue, stress and sleep indicate better response; High scores for general health, appetite, energy, anxiety, bowel habit, happiness indicate better response; See text for details | | | | | | |

**Supplementary File S3: Additional data for randomization baseline – standard of Care (SOC) drug dosage and site specific for SOC drug use and other study timelines**

**Table S3.1:** **Drugs with dosages used in the Standard of Care (SOC) treatment: a randomized controlled study to evaluate the co-administration of AYUSH-64 with Standard of Care (SOC) in mild - moderate symptomatic COVID-19 .**

| SOC Medicine  (Tab: Tablet) | Dosage |
| --- | --- |
| Tab Azithromycin | 500 mg od x3-5 days |
| Tab Doxycycline | 100 mg bid x 5 days |
| Tab Hydroxychloroquine sulphate | 400 mg od x 5 days (1st day bid) |
| Tab Zincovit (Zinc plus Vitamin B complex) | 50-100 mg (zinc content( od |
| Tab Vitamin C | 500 mg bid |
| Tab Multivitamin | Composite |
| Tab Vitamin D3 | 400 iu od, (with calcium citrate 500 mg) |
| Tab Pantoprazole | 20-40 mg od |
| Tab Paracetamol | 500-650 mg ,1-2 times on need basis |
| Tab Cetrizine | 5-10 mg od x 3-5 days |
| Tab Ivermectin | 12 mg od X 5 days |
| Injection Dexamethasone | 4-8 mg i/v stat |
| Injection Low molecular weight heparin analogue (clexane) | 40 mg s/c od x5-10 days |
| Oxygen intermittent | 2-4 l/min, nasal face mask |

**Table S3.2: Site specific data on some clinical variables (timelines) and individual standard of care drugs use (number of study participants and proportion %) in a randomized controlled study to evaluate the co-administration of AYUSH-64 with Standard of Care (SOC) in mild - moderate symptomatic COVID-19 (See main text for details)**

|  | Mumbai (n=60) | Nagpur (n=30) | Lucknow (n=49) |
| --- | --- | --- | --- |
| **Onset-Symptom to RT-PCR assay (days, mean, standard deviation)** | | | |
| AYUSH plus | 7 ± 5.6 | 0.5 ± 1.5 | 4.3 ± 2.8 |
| Standard of Care | 6.2 ± 4.5 | 1.5 ± 3.3 | 4.8 ± 3.8 |
| **RT-PCR assay to Hospitalization (days, mean, standard deviation)** | | | |
| AYUSH plus | 2.5 ± 1.1 | 2.0 ± 0.1 | 3.0 ± 0.8 |
| Standard of Care | 2.6 ± 0.8 | 2.0 ± 0.1 | 3.2 ± 0.7 |
| **Hospitalization to randomization (days, mean, standard deviation)** | | | |
| AYUSH plus | 1.1 ± 0.6 | 1.3 ± 0.5 | 1.6 ± 1.1 |
| Standard of Care | 1.1 ± 0.4 | 1.5 ± 1.2 | 1.8 ± 0.9 |
| **Tablet Azithromycin** | | | |
| AYUSH plus | 30 (50) | 12 (40) | 6 (12.2) |
| Standard of Care | 28 (46.6) | 15 (50) | 6 (12.2) |
| **Tablet Hydroxychloroquine Sulphate** | | | |
| AYUSH plus | 14 (23.3) | 14 (46.6) | 1 (2.0) |
| Standard of Care | 08 (13.3) | 15 (50) | 1 (2.0) |
| **Zincovit (zinc plus Vitamin B complex) Tablet** | | | |
| AYUSH plus | 11 (18.3) | 12 (40) | 25 (51) |
| Standard of Care | 02 (3.3) | 15 (50) | 25 (51) |
| **Vitamin C Tablet** | | | |
| AYUSH plus | 30 (50) | 14 (46.6) | 25 (51) |
| Standard of Care | 29 (48.3) | 15 (50) | 25 (51) |
| **Tablet Pantoprazole** | | | |
| AYUSH plus | 28 (46.6) | 13 (43.3) | 25 (51) |
| Standard of Care | 26 (43.3) | 14 (46.6) | 25 (51) |
| **Tablet Paracetamol** | | | |
| AYUSH plus | 19 (31.6) | 15 (50) | 25 (51) |
| Standard of Care | 15 (25) | 15 (50) | 25 (51) |
| **Tablet Cetrizine** | | | |
| AYUSH plus | 0 | 13 (43.3) | 0 |
| Standard of Care | 1 (1.6) | 14 (46.6) | 0 |
| **Intermittent Oxygen** | | | |
| AYUSH plus | 9 (15) | 0 | 0 |
| Standard of Care | 5 (8.3) | 1 (3.3) | 0 |

**Supplementary File S4: Adverse Events**

**Table S 4.1: Number of Adverse Events at all endpoints by System organ classification and Preferred term in a randomized controlled study to evaluate the co-administration of AYUSH-64 with Standard of Care (SOC) in mild - moderate symptomatic COVID-19 (n=139)**

|  |  | SOC (n=70) | | | | AYUSH plus (n=69) | | | |
| --- | --- | --- | --- | --- | --- | --- | --- | --- | --- |
| System organ classification (WHO) | Preferred term | During Hosp. | Week 4 | Week 8 | Week 12 | During Hosp | Week 4 | Week 8 | Week 12 |
| Cardiac | Raised Blood Pressure | - | - | - | - | - | 1 | - | - |
| Ear and labyrinth | Ear Ache | - | - | - | 1 | - | - | - | - |
| Gastrointestinal | Gastritis | - | 1 | - | - | - | - | - | - |
|  | Abdominal Discomfort | - | - | 1 | - | - | 1 | - | - |
|  | Diarrhea | - | 1 | - | - | - | 3 | 1 | - |
|  | Constipation | - | - | - | 1 | - | - | 1 | 1 |
|  | Epigastric Pain | - | - | - | - | - | - | 1 | - |
|  | Hyperacidity | - | - | - | - | - | 1 | 1 | - |
|  | Abdominal Pain | - | 1 | - | - | - | - | - | - |
| Hepatobiliary | Raised SGOT/SGPT | - | - | - | 1 | - | - | - | - |
| Infections and infestations | Fever | - | 1 | - | 1 | - | 1 | - | 4 |
|  | Malaria (P Vivax) | 1 | - | - | 1 | - | - | - | - |
|  | Cellulitis | 1 | - | - | - | - | - | - | - |
|  | Sore throat | - | 2 | - | 2 | - | 1 | - | - |
| Musculoskeletal and connective tissue | Neck Pain | - | - | - | 1 | - | - | 1 | 1 |
|  | Backache | - | - | - | 1 | 1 | - | - | - |
|  | Leg Pain | - | - | - | - | - | - | - | 1 |
|  | Ankle Pain | - | - | - | - | - | - | - | 1 |
|  | Joint Pain | - | - | - | - | - | 1 | - | - |
| Skin and subcutaneous tissue | White patches hands | - | - | 1 | - | - | - | - | - |
|  | Itching | - | - | - | - | - | - | 1 | - |
|  | Eczema | - | - | - | - | - | 1 | - | - |
| Respiratory, thoracic and mediastinal |  |  |  |  |  |  |  |  |  |
|  | Cough | - | 1 | 1 | - | - | 1 | - | - |
|  | Breathlessness | - | 1 | 2 | 2 | - | 4 | 2 | - |
|  | Loss of smell + Loss of Taste + Sore Throat + Breathlessness | - | 1 | - | - | - | - | - | - |
| Nervous system | GB Syndrome | - | - | - | - | 1 | - | - | - |
|  | Vertigo | - | - | - | - | 1 | 1 | - | - |
| Renal and urinary | Burning Micturition | - | - | 1 | - | - | - | - | - |
| Endocrine | High Blood Glucose levels | - | 3 | - | 3 | - | 2 | - | 4 |
| Investigations | Increased Triglycerides, LDL | - | 1 | - | - | - | - | - | - |
| Others | Weakness | - | 4 | 2 | - | - | 1 | - | 1 |
|  | Chills | - | - | - | 1 | - | - | - | - |
|  | Myalgia | - | 2 | 2 | 2 | - | 1 | 1 | - |
|  | Headache | - | - | - | - | - | 1 | 2 | - |
| Total |  | 2 | 19 | 10 | 17 | 3 | 21 | 11 | 13 |
| Note: Hosp:hospitalization; Note: AYUSH plus: AYUSH 64+ SOC: n: number of participants; SGOT/SGPT: serum glutamaseoxalacetate, serum gluatamase; No AE recorded for disorders of blood and lymphatic, immune system, metabolism and nutrition, psychiatric, reproductive system and breast, eye, vascular system, congenital familial and genetic, injury poisoning and procedural complications, and surgical and medical procedures; See text for detail | | | | | | | | | |

**Supplementary File S5: Routine laboratory investigations**

**Table S 5.1: Laboratory investigations [Data expressed as Mean ± SD] in a randomized controlled study to evaluate the co-administration of AYUSH-64 with Standard of Care (SOC) in mild - moderate symptomatic COVID-19: per protocol completer analysis**

| ***Blood /serum assay*** | | | | | |
| --- | --- | --- | --- | --- | --- |
| **Haemoglobin (g/dl)** | | | | | |
| Timepoints | Baseline (n=139) | On Discharge  (n=137) | Week 4  (n=129) | Week 8  (n=127) | Week 12  (n= 120) |
| AYUSH plus | 13.62 ± 1.42 | 13.34 ± 1.49 | 13.18 ± 1.62 | 13.54 ± 1.61 | 13.53 ± 1.78 |
| SOC | 13.80 ± 1.62 | 13.41 ± 1.90 | 13.67 ± 1.29 | 13.80 ± 1.45 | 13.94 ± 1.39 |
| **Red Blood Cells – Total (million/mm^3^)** | | | | | |
| AYUSH plus | 4.71 ± 0.59 | 4.69 ± 0.54 | 4.56 ± 0.55 | 4.65 ± 0.56 | 4.77 ± 0.63 |
| SOC | 4.69 ± 0.68 | 4.81 ± 0.70 | 4.64 ± 0.52 | 4.71 ± 0.55 | 4.81 ± 0.54 |
| **White Blood Cells – Total (cells/ mm^3^)** | | | | | |
| AYUSH plus | 5920.69 ± 2008.8 | 6781.39 ± 1513.6 | 7156.6 ± 1634.0 | 7228.57 ± 1334.6 | 6827.27 ± 1747.7 |
| SOC | 6828.30 ± 2085.8 | 6650 ± 1911.0 | 6888 ± 1426.7 | 7024.49 ± 1292.2 | 7001.92 ± 1450.8 |
| **Platelets (lakhs/ mm^3^)** | | | | | |
| AYUSH plus | 2.73 ± 1.31 | 3.88 ± 1.63 | 2.60 ± 0.98 | 2.60 ± 0.87 | 2.63 ± 0.86 |
| SOC | 2.55 ± 1.48 | 3.34 ± 1.55 | 2.39 ± 1.00 | 2.33 ± 0.93 | 2.48 ± 0.93 |
| **Erythrocyte sedimentation rate** **(mm/hour)** | | | | | |
| AYUSH plus | 50.19 ± 38.03 | 42.98 ± 36.22 | 31.63± 26.21 | 26.77± 22.36 | 21.10 ± 21.05 |
| SOC | 46.86 ± 37.37 | 42.68 ± 34.19 | 21.07± 14.27 | 23.77± 15.10 | 15.75 ± 10.76 |
| ***Liver Function Tests*** | | | | | |
| **Total Bilirubin** (mg/dL) | | | | | |
| AYUSH plus | 0.64 ± 0.28 | 0.61 ± 0.27 | 0.70 ± 0.30 | 0.75 ± 0.47 | 0.79 ± 0.37 |
| SOC | 0.78 ± 0.51 | 0.64 ± 0.35 | 0.80 ± 0.34 | 0.82 ± 0.35 | 0.88 ± 0.39 |
| **Total Proteins** (mg/mL) | | | | | |
| AYUSH plus | 7.12 ± 0.59 | 8.33 ± 3.42 | 7.20 ± 0.51 | 7.23 ± 0.57 | 7.34 ± 0.67 |
| SOC | 7.09 ± 0.66 | 7.07 ± 0.57 | 7.20 ± 0.52 | 7.16 ± 0.52 | 7.16 ± 0.74 |
| **Serum Globulin** (g/dL) | | | | | |
| AYUSH plus | 3.14 ± 0.49 | 3.20 ± 0.35 | 3.12 ± 0.39 | 3.14 ± 0.39 | 3.10 ± 0.44 |
| SOC | 3.08 ± 0.61 | 3.13 ± 0.52 | 3.07 ± 0.40 | 3.08 ± 0.39 | 2.99 ± 0.50 |
| **Serum Albumin** (g/dL) | | | | | |
| AYUSH plus | 3.99 ± 0.39 | 3.89 ± 0.35 | 4.07 ± 0.30 | 4.09 ± 0.28 | 4.24 ± 0.38 |
| SOC | 4.0 ± 0.41 | 3.89 ± 0.41 | 4.14 ± 0.31 | 4.08 ± 0.23 | 4.15 ± 0.39 |
| **Albumin/Globulin ratio** | | | | | |
| AYUSH plus | 1.34 ± 0.38 | 1.23 ± 0.19 | 1.32 ± 0.20 | 1.32 ± 0.17 | 1.38 ± 0.20 |
| SOC | 1.36 ± 0.33 | 1.31 ± 0.24 | 1.47 ± 0.59 | 1.36 ± 0.18 | 1.40 ± 0.21 |
| **Aspartate aminotransferase** (units/L) | | | | | |
| AYUSH plus | 42.96 ± 32.33 | 36.91 ± 28.39 | 28.64± 14.29 | 27.48± 14.02 | 30.44 ± 18.15 |
| SOC | 43.93 ± 28.49 | 37.99 ± 24.78 | 33.26± 22.32 | 29.47± 11.35 | 31.41 ± 18.79 |
| **Alkaline Phosphatase** (units/L) | | | | | |
| AYUSH plus | 106.45 ± 63.37 | 85.33 ± 40.38 | 103.78± 45.26 | 116.93± 66.48 | 108.38 ± 53.72 |
| SOC | 113.80 ± 68.58 | 91.25 ± 53.91 | 111.29 ± 51.17 | 116.78 ± 63.57 | 110.37 ± 61.62 |
| **Renal function test** | | | | | |
| **Serum Creatinine** (mg/dL) | | | | | |
| AYUSH plus | 1.02 ± 0.23 | 0.96 ± 0.50 | 0.99 ± 0.23 | 1.02 ± 0.23 | 1.01 ± 0.24 |
| SOC | 1.00 ± 0.31 | 1.08 ± 0.68 | 0.92 ± 0.22 | 0.93 ± 0.19 | 1 ± 0.29 |
| **Blood Urea Nitrogen** (mg/dL) | | | | | |
| AYUSH plus | 14.97 ± 8.28 | 10.74 ± 7.25 | 14.53± 12.17 | 17.09± 13.09 | 15.70 ± 10.16 |
|  | 17.25 ± 10.82 | 10.91 ± 7.05 | 15.97± 10.02 | 17.43± 10.50 | 17.02 ± 9.86 |
| **Blood Sugar Level** (mg/dL) | | | | | |
| AYUSH plus | 112.50 ± 37.54 | 120.11 ± 54.21 | 124.51 ± 59.30 | 96.61± 18.49 | 98.97 ±15.40 |
| SOC | 114.17 ± 35.23 | 125.37±34.90 | 122.39± 58.25 | 92.99± 10.83 | 92.26±11.09 |
| Note: AYUSH plus: AYUSH 64 + SOC; No significant difference between the groups for any of the assay shown above at significant p<0.05, ANOVA; n; number of participants; study patients with diabetes excluded for blood sugar data shown above. See text for detail. | | | | | |
